# Effectiveness of the PAS Scale for Diagnosing the Severity of Acute Appendicitis in Children: A Cohort Study

**DOI:** 10.1101/2024.12.07.24318625

**Authors:** Sucso-Noa Edgar

## Abstract

**What’s Known on This Subject:** Appendicitis is an acute inflammatory process that affects the appendix, one of the common causes of abdominal pain in children in emergency. Being one of the most common conditions faced by surgeons.

The PAS scale, a clinical score for acute appendicitis, is among the most popular for use in children for diagnosis. The severity of appendicitis subclassified as simple appendicitis (congested or suppurated) vs complicated appendicitis (gangrenous or perforated) could be predicted with a PAS scale ≥8.

**What This Study Adds:** Tests logistic regression models as a basis for developing more complex models to improve prediction of complicated acute appendicitis in children.

**Background:** The Pediatric Appendicitis Score (PAS) is a highly utilized diagnostic tool for acute appendicitis in pediatric patients. The severity of appendicitis subclassified as simple appendicitis (congested or suppurated) vs complicated appendicitis (gangrenous or perforated) could be predicted with a PAS scale ≥8.

**Methods:** The type of study of the present research, according to Altman Douglas, is: Observational, Retrospective y Longitudinal. The design is cohort-type. The population was evaluated, 86 children aged 4-14 years with a preoperative diagnosis of appendicitis, grouped into 2 groups: complicated appendicitis (43) and simple appendicitis (43) exposed to the PAS≥8 or PAS 8.

**Results:** The effectiveness of the PAS≥8 scale for diagnosing the severity of appendicitis showed a predictive diagnostic accuracy of 59.3% and increases the likelihood of severity by 2.246 times (CI:95% 0.917-5.50 p=0.077) in the predictive model. There were statistically significant differences in cough/hop/percussion Tenderness, migration of pain, anorexia, leukocytosis and neutrophilia, between scale PAS≥8 vs PAS<8.

**Conclusion:** The PAS≥8 scale alone is not sufficient to diagnose the severity of acute appendicitis, with 59.3% predictive diagnostic accuracy and increasing the likelihood of presenting the severity of appendicitis by 2.246 times.

## INTRODUCTION

Acute appendicitis is a common cause of acute abdomen in emergencies. Its progression can become complicated if not diagnosed early or if its severity is not anticipated. Globally, the incidence of acute appendicitis is 100 new cases per 100 000 people per year(1). In the U.S. 70 000 pediatric appendectomies are performed annually additionally, in the age group of 5 to 11 years, the incidence reaches 36%, with an average of 1.38 cases per 1,000 children(2,3).

For the diagnosis of appendicitis, clinical findings and auxiliary examinations are required. In children, it is not easy to determine the severity of the condition.This could be improved by using techniques such as the Pediatric Appendicitis Score (PAS) in emergency care.

Recently, the World Society of Emergency Surgery in their Jerusalem Guidelines reached a consensus that the PAS scale is a useful and sensitive tool to exclude acute appendicitis and recommended not making the diagnosis based solely on clinical scales in those with suspected pediatric appendicitis(4).

The PAS scale developed by Madan Samuel is still relevant and applicable today. It consists of 8 parameters, with the main ones being (**T**enderness in the right lower quadrant and cough/hop/percussion **T**enderness) and the other secondary parameters (**M**igration of pain, **A**norexia, **N**ausea/vomiting, **E**levated temperature, **L**eukocytosis, and **N**eutrophilia) **TT/MANELN**, being useful for predicting the risk of pediatric appendicitis(5).

The effectiveness of the PAS scale for diagnosing the severity of appendicitis is defined as a mechanism to achieve predictive diagnostic accuracy of the PAS scale≥8 for severity and predictive possibility through the binomial logistic regression model, for the number of correct cases over a period of 1 year and 9 months.

In a study of 72 patients in a hospital in Japan, it suggests that the PAS scale would have a correlation with the severity of appendicitis because they found with greater complications and prolonged hospital stay than those with PAS[8(6).

The study aims to evaluate the effectiveness of the PAS≥8 scale in diagnosing the severity of acute appendicitis in children and as secondary objectives the PAS characteristics with respect to other variables.

## METHODO

### Study Type

The type of study of the present research, according to Altman Douglas (7), is: Observational, Retrospective, Longitudinal. The design is of the cohort type, to evaluate the effectiveness of the PAS scale in diagnosing the severity of appendicitis.

### Participans

The population consists of all patients aged 4 to 14 who are admitted with a diagnosis of acute appendicitis to the emergency department, undergo open appendectomy with intraoperative findings of simple or complicated appendicitis, and are hospitalized in the Pediatric Service from the Carlos Monge Medrano Hospital during 2020 to 2022, which meet selection criteria.

The 86 patients who met the criteria detailed in the inclusion flow, for more details see (results section). He grouped into two groups: First group, of 43 children with Simple Appendicitis (the surgical findings of: Congestive Appendicitis and Suppurative Appendicitis were considered). A second group, consisting of 43 children with Complicated Appendicitis (the surgical findings of Necrotizing Appendicitis and Perforated Appendicitis were considered). Both groups were exposed to PAS≥8 or PAS 8.

### Variables

#### Severity of appendicitis

The severity of appendicitis in the present research is referred to as the differentiation between complicated appendicitis and simple appendicitis based on the intraoperative findings discovered by the surgeon during the open appendectomy.

### Simple Appendicitis

It is an early phase appendicitis that includes congestive (catarrhal) appendicitis and suppurative (phlegmonous) appendicitis in the intraoperative findings. This type of appendicitis has not yet reached the stage of complications.

### Complicated Appendicitis

It is a perforated appendicitis as a common component in addition to gangrene, pus, purulent peritonitis, presence of a fecalith or abscess(4). Complicated appendicitis includes: necrotizing (gangrenous) appendicitis, due to the microperforations observed, and perforated appendicitis found in intraoperative findings, in some cases with: localized peritonitis, generalized peritonitis, and abscesses.

#### PAS scale

The 8 parameters(TT/MANELN): Main parameters (**T**enderness in right iliac fossa when coughing/jumping/percussing, manifests during the patient’s physical examination; **T**enderness in the right lower quadrant is a symptom that the patient exhibits at the level of the right iliac fossa, most representative in the late stages of appendicitis) and secondary parameters (**M**igration of pain is when abdominal pain changes position from being periumbilical or diffuse to localizing in the right lower quadrant, **A**norexia, it is the decrease in appetite, an early manifestation may or may not be present, **N**ausea/vomiting, it is when the patient expresses a nauseous feeling alone or it may be followed by vomiting, **E**levation of temperature, for this research we consider T≥37.5 Grades Celsius(°C), it is a thermal rise sensation quantified at the axillary level, **L**eukocytosis is the value above >10,000/μL of leukocytes, a range that exceeds normal, **N**eutrophilia is considered an absolute neutrophil count value >7.5 000/μ.).

The PAS≥8 and PAS<8 scales were evaluated upon admission to the emergency room and follow-up was conducted because all were exposed to various PAS scale scores. Additionally, regarding its severity, the severity variable in emergency simple appendicitis vs complicated appendicitis was also evaluated, and after the surgical finding, simple appendicitis vs complicated appendicitis was also verified.

### Data analysis

The statistical software Excel was used to analyze the database without selection criteria and another with selection criteria, and then it was processed in the statistical software Jamovi 2.3.28 (8).

Descriptive statistics were used for: frequencies, means, SD, medians, minimums, and maximums for the analysis of the variables. Additionally, binomial logistic regression was used for predictive diagnostic accuracy of severity (through diagnostic accuracy of the ROC curve (AUC)) and predictive possibility (OR) in the predictive model for the PAS scale with a 95% CI.

Assisted by ROC curves for sensitivity, specificity, area under the ROC curve, PPV, NPV, and OR measures. The highest Youden index was used to determine the cutoff value of the score on the PAS scale using the Friesen Plugin, PPDA (ROC Test) for Jamovi.

We used Jamovi for Shapiro Wilk, Whitney U-test and frequencies (contingency table) was used for quantitative variables for independent samples for the present study, statistical significance is p<0.05.

#### Ethical aspects

It was a secondary data study and there was no contact with the patient, therefore patient consent was not requested. The patient’s anonymity and the confidentiality of data such as identity or any other information that could compromise the patient were maintained.

## RESULTS

Between January 2020 and September 2022, 86 children were studied at HCMM using a cohort design. Eligibility criteria were applied, with stages of evaluation, inclusion, and exclusion, as well as losses during follow-up, until the process was completed as detailed in (Figure 1).

The losses during the follow-up were due to: 1 did not have a report of findings and the remaining 24 had two to three findings of appendicitis (Example: Necrotizing appendicitis + perforated appendicitis or suppurative appendicitis + necrotizing appendicitis + perforated appendicitis/appendiceal abscess).

### Characteristics of the participants

The mean age of patients was 9.6 ±3 years, Male 53.5% and similar distribution of rural and urban(p=0.982) in patients with PAS ≥8 and PAS <8. There were statistically significant differences in the cough/hop/percussion Tenderness (100% vs 78.1% p<0.001), migration of pain (77.8% vs 31.3% p<0.001), anorexia (42.6% vs 12.5% p=0.004), leukocytosis (96.3% vs 62.5% p<0.001) and neutrophilia (100% vs 65.6% p<0.001) between scale PAS≥8 vs PAS<8. Tenderness right lower Quadrant (RLQ) found in almost all patients (98.8% p=0.191) in the mnemotechnic **TT/MANELN** (Table 1) and Appendicitis Complicated were more common in PAS ≥8 compared a PAS <8(57.4% vs 37.5% p=0.074) (Table 2).

**Table 1.** Scale PAS (TT/MANELN Mnemotechnic).

| Parameters |  | Score |
| --- | --- | --- |
| Main Parameters | **T**enderness in right lower quadrant | 2 |
|  | Cough/Hop/Percussion **T**enderness | 2 |
| Secondary Parameters | **M**igration of pain | 1 |
|  | **A**norexia | 1 |
|  | **N**ausea/vomiting | 1 |
|  | **E**levated temperature | 1 |
|  | **L**eucocytosis | 1 |
|  | **N**eutrophilia | 1 |

Based in: Samuel M. Pediatric appendicitis score. J Pediatr Surg.2002

**Table 2.**
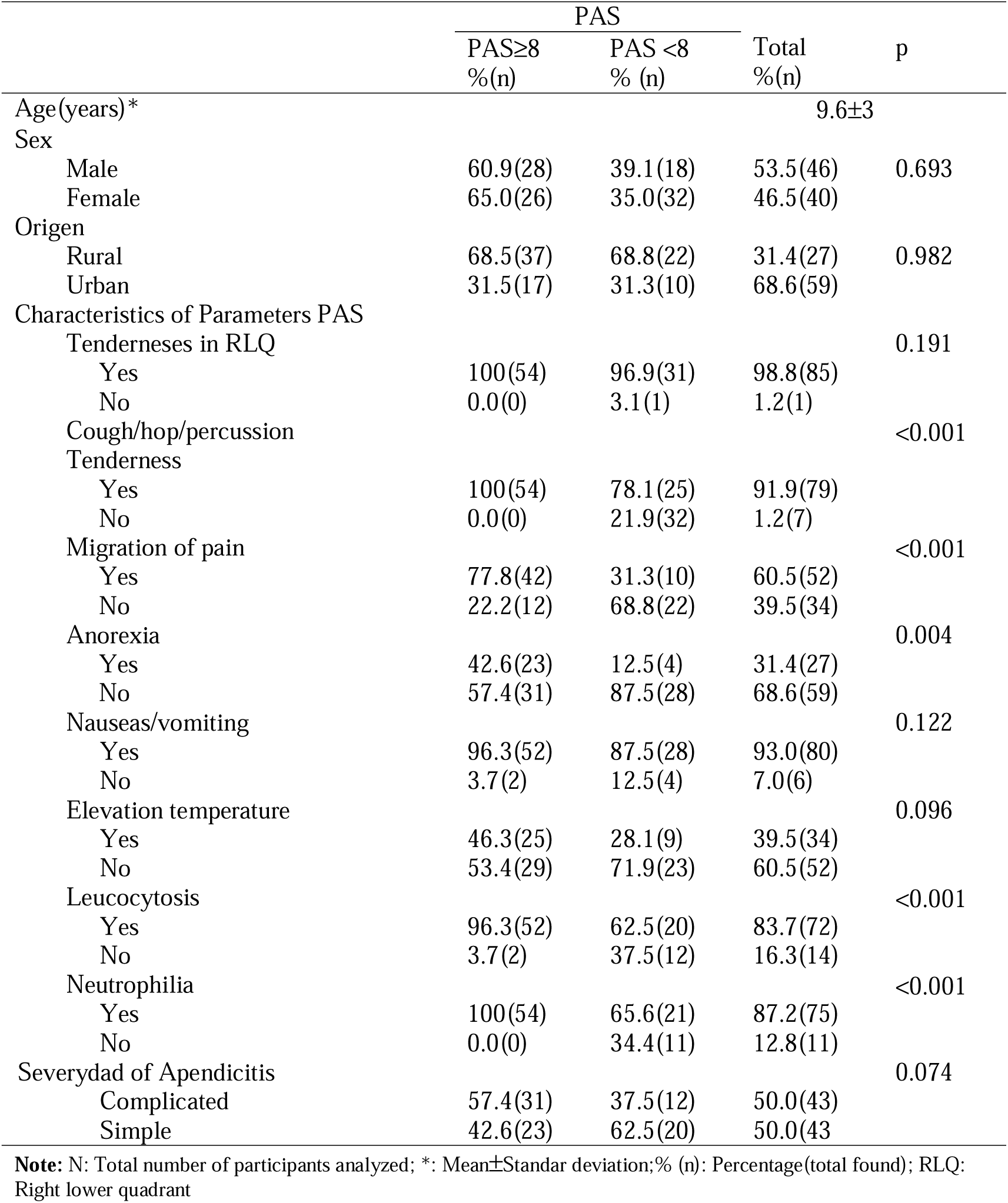
Characteristics of the patients in the study(N=86).

#### Analysis for the main objective

The effectiveness of the PAS≥8 scale showed a diagnostic accuracy of 59.3% to predict the severity of appendicitis in the binomial logistic regression model, a cutoff value of 0.5. For the PAS≥8 scale as a predictor for diagnosing the severity of acute appendicitis, an ROC curve was designed in which the sensitivity was found to be 72.1%, the specificity was 46.5%, and the area under the curve was 0.593 for the model. (Figure 2). Obtaining a PAS≥8 score increases the likelihood of presenting with severe appendicitis by 2.246 times compared to those with a PAS<8 score (CI: 95% 0.917 to 5.50 p=0.077), a statistically non-significant result, to see (Table 3).

**Figure 2.**
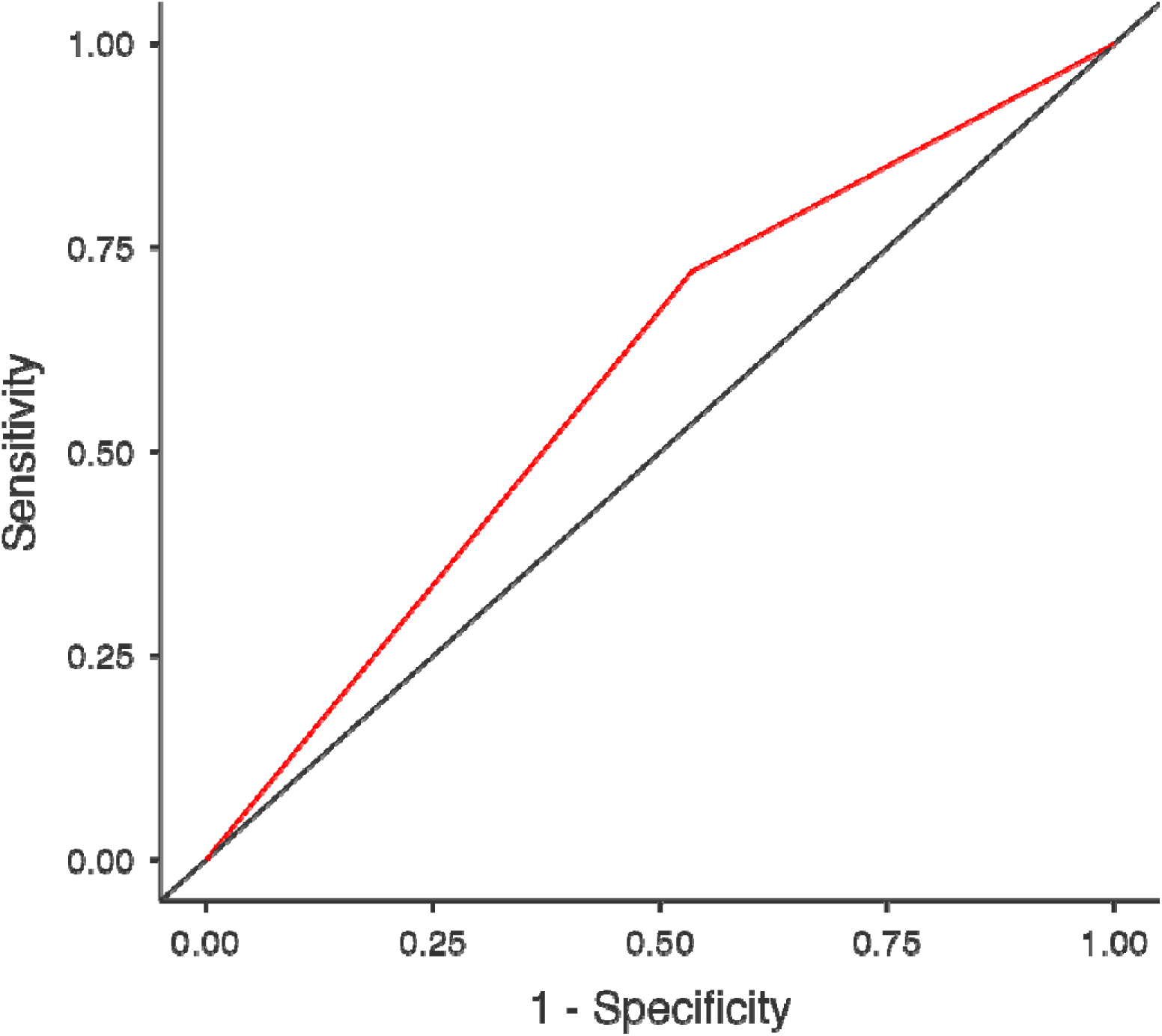
ROC Curve.

**Table 3.** Model Coefficients-Severity of Acute Appendicitis(N=86).

| Predictor | Estimate | SE | Z | p | OR | 95% Confidence Interval |  |
| --- | --- | --- | --- | --- | --- | --- | --- |
|  |  |  |  |  |  | Lower | Upper |
| Intercept | -0.511 | 0.365 | -1.40 | 0.162 | 0.600 | 0.293 | 1.23 |
| PAS≥8: | | | | | | | |
| Si – No | 0.809 | 0.457 | 1.77 | 0.077 | 2.246 | 0.917 | 5.50 |

Note. Estimates represent the log odds of "Severity= Complicated Appendicitis " vs. "Severity= Simple Appendicitis " ; N: Total number of participants analyzed; OR: **Odds ratio**

The logistic regression model is employed in clinical studies with the following formula.(9):

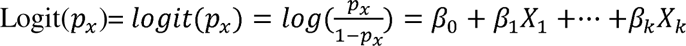

The formula, to assess the effectiveness of the probability of predicting the effectiveness of the PAS≥8 scale for diagnosing the severity of appendicitis is:

Probability of predicting severity = −0.511 + 0.809×PAS≥8 **R^2^_McF=_**0.0269

**Additional approaches to the treatment of appendicitis include:**

Garcia-Amado C. et al. (10):

Probability predicting = t = -(−9.99 + 0.030xage(years) + 0.016xduration of symptons(h) + 0.084xpercentage of neutrophils (%) + 0,008xCRP(mg/L))

Feng W. et al.(11):

Probability predicting=u= -(2.997-1.559xage(years) + 0.090x white blood cell count(WBC)(10^9^/L)+0.010xDuration of symtoms(hours))

Eddama M. et al.(12):

Probability predicting =v = -(−8.814 + 0.364xlog 2 CRP + 1.768xlog 2 WWC + 0.025xage + 0.647x(0 if Female/ 1 if Male)

Chambers A.C. et al(13)

Probability predicting = w = -(−2.77+0.005xCRP + 0.061xBilirubin + 0.211xWCC)

### Analysis for secondary objectives

In our study, no cases of PAS<5 and we were observed Positive and negative predictive values for each score (Table 4).In the evaluated patients, there were statistically significant: leukocytosis (16.98×10^3/µL ±4.81 vs 13.29×10^3/µL ±5.69 p[0.001), neutrophilia(14.70 x10^3/µL ±4.68 vs 11.06 x10^3/µL ±4.68 p[0.001), PAS score (8.59±0.59 vs 6.38±0.71 p[0.001), between scale PAS≥8 vs PAS<8 (Table 5).

**Table 4.** Positive and negative predictive values for each score.

| PAS | Sensitivity (%) | Specificity (%) | PPV (%) | NPV (%) | Youden's index | AUC | Metric Score |
| --- | --- | --- | --- | --- | --- | --- | --- |
| 1 | 0 | 0 | 0 | 0 | 0 | 0 | 0 |
| 2 | 0 | 0 | 0 | 0 | 0 | 0 | 0 |
| 3 | 0 | 0 | 0 | 0 | 0 | 0 | 0 |
| 4 | 0 | 0 | 0 | 0 | 0 | 0 | 0 |
| 5 | 100% | 0% | 50% | 0% | 0.00 | 0.58 | 1.00 |
| 6 | 90.7% | 0% | 47.56% | 0% | -0.09 | 0.58 | 0.90 |
| 7 | 86.05% | 23.26% | 52.86% | 62.5% | 0.09 | 0.58 | 1.09 |
| 8 | 72.09% | 46.51% | 57.41% | 62.5% | 0.19 | 0.58 | 1.18 |
| 9 | 37.21% | 69.77% | 55.17% | 52.63% | 0.07 | 0.58 | 1.07 |
| 10 | 6.98% | 100% | 100% | 51.81% | 0.07 | 0.58 | 1.07 |

Note: PAS: Pediatric Appendicitis Score, PPV: Positive Predictive Value, NPV: Negative Predictive Value; AUC: Receiver Operating Characteristic

**Table 5.** Description of leukocytosis, neutrophilia, temperature elevation, segmented neutropils, band neutrophils and score PAS and scale PAS≥8.

| | PAS≥8 | N | Mean | SD | p Mann<br>Wiltney | p Shapiro-Wilk |
| --- | --- | --- | --- | --- | --- | --- |
| Hospital Stay | Si | 54 | 6.17 | 2.081 | 0.116 | <□0.001 |
| | No | 32 | 5.81 | 2.912 | | <□0.001 |
| Leukocytosis<br>x10<sup>3</sup>/μL | Si | 54 | 16.98 | 4.962 | <□0.001 | 0.066 |
|  | No | 32 | 13.29 | 5.693 |  | 0.003 |
| Neutrophilia<br>x10<sup>3</sup>/μL | Si | 54 | 14.70 | 4.678 | <□0.001 | 0.038 |
|  | No | 32 | 11.06 | 5.562 |  | 0.011 |
| Score PAS | Si | 54 | 8.59 | 0.599 | <□0.001 | <□0.001 |
| | No | 32 | 6.38 | 0.707 | | <□0.001 |
| Temperature elevation | Si | 54 | 37.22 | 0.746 | 0.697 | <□0.001 |
|  | No | 32 | 37.19 | 0.898 |  | 0.015 |
| Segmented neutropils | Si | 54 | 84.80 | 6.363 | 0.008 | <□0.001 |
|  | No | 32 | 79.22 | 10.646 |  | 0.002 |
| Band neutropils | Si | 54 | 1.38 | 2.244 | 0.406 | <0□.001 |
| | No | 32 | 1.82 | 2.690 | | <□0.001 |

SD: Standar deviation; Number of participants analyzed=86; N: Total

We obtained higher percentages in the PAS≥8 scale: duration of illness(24 to 48h 33.7% p=0.025),perforated appendicitis(31.4%p[0.001), Rockey Davis incision (33.7% p=0.004)and appendicitis with generalized peritonitis(19.8% p[0.001) all of the above when compared to the PAS[8(Table 6).

**Table 6.**
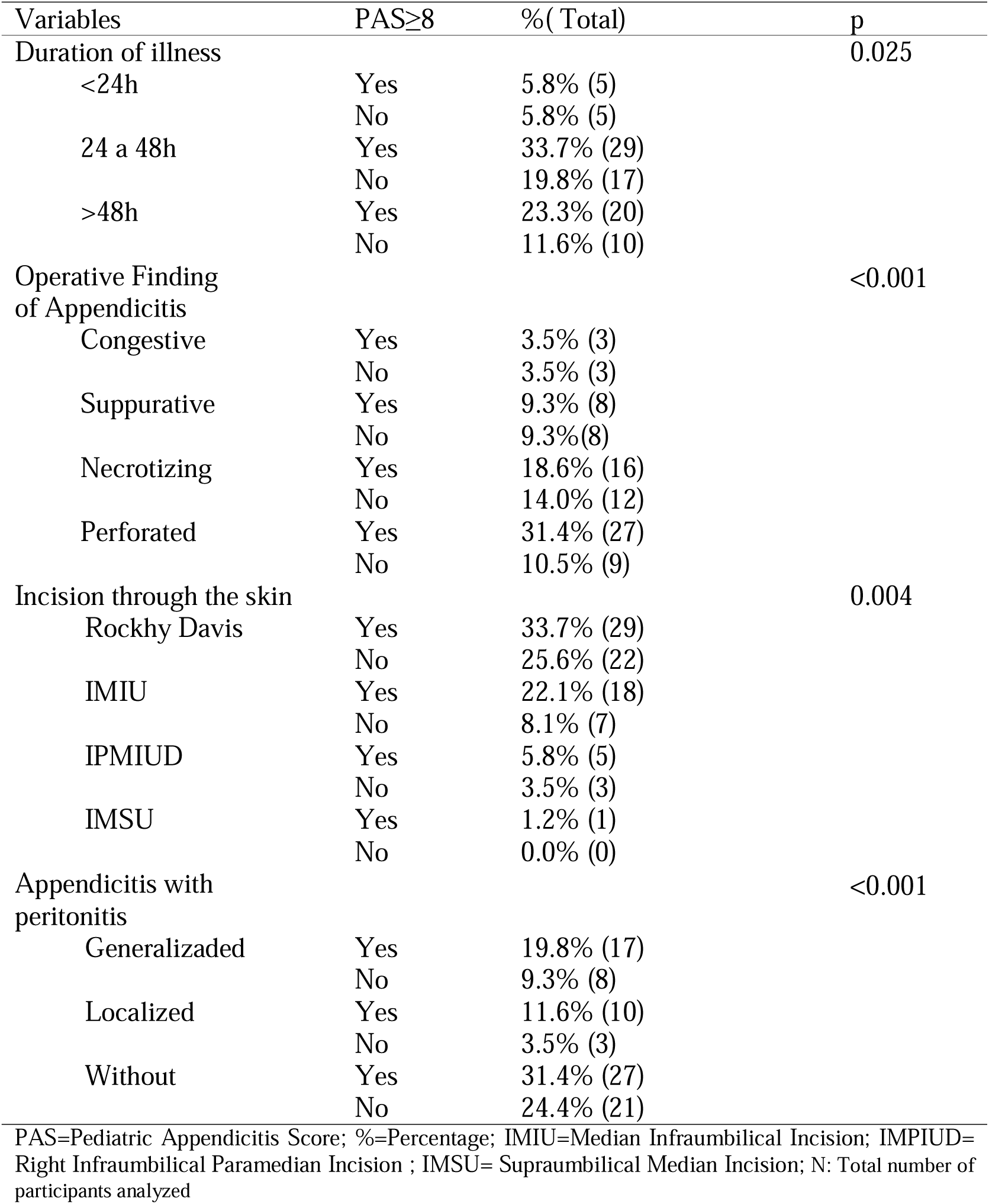
Characteristics of the duration of duration of illness and operative findings compared with a PAS≥8(N=86).

## DISCUSSION

In the latest update of the Jerusalem Guidelines by the World Society for Emergency Surgery, the PAS Scale is considered one of the most used clinical scoring systems in children(4). Current scoring systems (PAS, Lintula, Alvarado, MPAS and Tzanakis) help us in the diagnosis of appendicitis and reduce negative appendectomy rates in children at present(14).

Our study was to evaluate the effectiveness for diagnosing the severity of acute appendicitis (complicated appendicitis and simple appendicitis) using the PAS scale≥8.The results of the cut-off value of the PAS score equal to 8 in the PAS scale to diagnose the severity of appendicitis agree with the study of Fugii et al.(6).

PAS, in conjunction with symptom duration, may assist in predicting patients with a higher likelihood of developing a postoperative intraabdominal abscess(15).PAS≥8 scale only was not effective, for diagnosing complicated appendicitis.

In our study, no cases of PAS<5 were observed; we agree with another study that did not observe PAS<4(16).

A study in a hospital, analyzed in 161 children three predictors: the PAS≥8 scale, CRP>4mg/dl and symptom duration>1day for complicated appendicitis, they designed a ROC curve for the three predictors obtaining: an area under the curve 0.91, sensitivity of 51%, specificity of 99%, PPV of 83% and NPV of 66%, different from our study that only analyzed one predictor which was the PAS≥8 scale(17).Higher CRP levels and PAS were associated with increased histologic inflammation of the appendix(18).

Currently, scoring systems based on NLR(NLR: neutrophil-to-lymphocyte ratio), PLR(platelet-to-lymphocyte ratio), and LMR(lymphocyte-to-monocyte ratio) reference values vary according to age and gender(19). New regression analyses could include the PAS scale to distinguish complicated appendicitis and simple appendicitis in children. As a scoring system called POPs, which combines inflammatory predictors, ultrasound findings(20). The clinical prediction rules, which combine clinical and objective variables, had the highest discriminant capacity(21).

In addition, another study of 260 children evaluated the performance of the PAS scale and found an area under the curve of 0.992, sensitivity of 98.74%, specificity of 95.65%, PPV of 95.7%, and NPV of 96.65% for a PAS≥6(22). In 104 children studied, sensitivity of 96.8%, specificity of 80%, PPV of 98.91%, NPV of 57.14% and area under the curve of 0.84(23). Both studies contradict ours because they are for the diagnosis of appendicitis but not for the severity of acute appendicitis.

The most accurate predictors of appendicitis (both simple and perforated) were rebound tenderness, hop/cough tenderness, laboratory results, and ultrasound findings, as demonstrated in a study of multi-center cohorts(24). Pediatric predictors could include neutrophilia, leukocytosis, pain upon cough/hop/percussion Tenderness, migration of pain, anorexia, as they are significant in the present investigation or the PAS≥8 scale to be evaluated alongside other predictive models. In another study PAS score ≥ 7 is associated with prolonged hospital stay(25).

A study of 1141 children showed that delaying appendectomy within 24h after the onset of appendicitis is safe and feasible(26). The duration of illness between 24 and 48 hours is associated with a PAS≥8 scale.

The present research, during periods when open surgery was the only option at the hospital, the limitation was that being a retrospective study, there was no control of variables such as surgical findings.

The results obtained serve to think in regression models could help developing countries that include clinical variables and variables of basic laboratory packages due to the deficit of human resources and help with the diagnosis of the severity of appendicitis in children.

### Conclusion

The PAS≥8 scale alone is not sufficient to diagnose the severity of acute appendicitis, with a 59.3% predictive diagnostic accuracy and increasing the probability of presenting with the severity of appendicitis by 2.246 times. It could be combined PAS scale with other variables to create models that help differentiate complicated appendicitis and simple appendicitis in children.

## Acknowledgments

The author expresses gratitude to National University of the Altiplano for enabling the completion of this work as a requirement to obtain the bachelor’s degree in medicine. I would also like to thank the following individuals for guiding me in this work: Eng. Fred Torres Cruz, Dr. Francisco Armando Lajo Soto, Dr. Juan Cruz Cruz, Dr. Luis Alberto Villalta Rojas, Dr. Enrique Alfredo Capio, and Dr. Alejandro Vela Quico.

## Data Availability

To request access, for further research purposes, please to write to the corresponding author.

## Abbreviations

PAS: Pediatric appendicitis score
IY: Youden Index
USA: United States
CRP: C Reactive Protein
ROC: Receiver Operating Characteristic
AUC: Area under the curve
PPV: Positive predictive value
NPV: Negative predictive value
PMN: Polymorphonuclear
CI: confidence interval
SD: standard deviation

